# Pregnancy pharmacoepidemiology: How often are key methodological elements reported in publications?

**DOI:** 10.1101/2021.05.04.21256602

**Authors:** Andrea V Margulis, Alison T Kawai, Mary S Anthony, Elena Rivero-Ferrer

## Abstract

**Purpose:** Publications are an important information source for clinicians, researchers, and patients. Key methodological elements must be reported for maximum transparency. We identified key methodological elements necessary for fully understanding pharmacoepidemiological research in pregnancy and quantified the proportion of studies that report these elements in a sample of publications.

**Methods:** Key methodological elements were identified from guidelines from regulatory agencies, literature, and subject-matter knowledge: source of information to determine pregnancy start; mother- or father-infant linkages (process, success rate); unit of analysis; and whether non-live births and fetuses with various anomalies were included in the study population.

We conducted a literature review for recent observational studies on medical product utilization or safety during pregnancy and estimated the prevalence of reporting these elements.

**Results:** Data were extracted from a random sample of 100 publications; 8% were published in epidemiology/pharmacoepidemiology journals; 85% were medical product–safety studies.

Of included publications, 43% reported the source for determining pregnancy start; 57% reported whether the study population included multifetal pregnancies; 39%, whether it included more than 1 pregnancy per woman; 27%, whether it included fetuses with chromosomal abnormalities; 60%, fetuses with major congenital malformations; and 93%, non-live births. Of the 20 studies with mother-infant linkage, 35% described the process; 21% reported the linkage success rate. Among studies with more than one pregnancy/offspring per woman, 22% reported methods addressing sibling correlation.

**Conclusions:** In this sample of pregnancy-related pharmacoepidemiology publications, completeness of reporting can be improved. A pregnancy-specific checklist would help to increase transparency in the dissemination of study results.

**Key points:**

▪ Publications on the utilization or safety of medical products in pregnancy (pregnancy pharmacoepidemiology) are fully understandable when all key methodological elements are presented to the reader.
▪ We identified 17 methodological elements from guidelines, literature, and subject-matter knowledge that we felt were crucial for understanding publications on pregnancy pharmacoepidemiology.
▪ In a random sample of 100 publications, completeness of reporting varied across elements, with almost perfect reporting on whether non-live births were included in the study population to a 21% completeness on mother-infant linkage success rate.
▪ A pregnancy-specific checklist would help to increase transparency in the dissemination of study results.

## 1 INTRODUCTION

Publications are an important source of information for clinicians, researchers, and patients seeking information on medical product utilization and safety in pregnancy (hereafter, “pregnancy pharmacoepidemiology”). In order for those publications to have their full impact, reporting of the methods have to be complete. However, features of study design that are specific to research on the utilization and safety of medical products in pregnancy are sometimes omitted from publications. Such key design elements include the source of information on pregnancy start date (needed to understand the precision of the study exposure window) and the exact composition of the study population (e.g., whether multifetal pregnancies, fetuses/infants with chromosomal abnormalities, or fetuses/infants with minor congenital malformations were included in the study population), which can affect the prevalence of some outcomes and potentially impact relative risk or prevalence ratio estimates. Poor reporting can be perceived to indicate poor study quality,^1^ thereby hampering the reader’s ability to assess the risk of bias. Also, the results of studies that do not report their key methodological attributes in a complete manner can be difficult to interpret. Missing or incomplete information can limit researchers’ ability to compare results across studies and can also impact the acceptance of and understanding by clinical, research, and patient audiences.

Thorough reporting in health-related research is currently considered an important characteristic of a well-written study report, as evidenced by the 457 guidelines on health research reporting listed in the EQUATOR Network’s library of reporting for health research.^2^ Documents in this library include guidelines for reporting on research on obstetrics/pregnancy and on pharmacoepidemiology, but not specifically on pregnancy pharmacoepidemiology.

In this review, we first identified key methodological elements that we deemed should be reported by studies on pregnancy pharmacoepidemiology, taking into consideration regulatory guidelines from the United States (US) Food and Drug Administration and the European Medicines Agency, relevant literature, and subject-matter knowledge. We then assessed the prevalence of reporting of those elements in a sample of publications.

## 2 METHODS

### 2.1 Key methodological elements in pregnancy pharmacoepidemiology

The authors reviewed the recently updated US and European regulatory guidelines for conducting observational studies on the pregnancy safety of medical products^3,4^and the recent literature and used their expertise to identify methodological elements that are considered key in research on pregnancy pharmacoepidemiology. When there was disagreement, authors discussed the issue until reaching consensus.

The research team initially identified a relatively extensive list of data elements. Some elements were later found to be duplicative or not informative. The final list consisted of 17 key elements organized in four domains: (1) source of information on start and end of pregnancy; (2) composition of the study population, including, as separate elements, whether multifetal pregnancies, more than one pregnancy per woman, fetuses with chromosomal abnormalities, fetuses with major or minor malformations, and non-live births were included in the study population; (3) mother-infant and father-infant linkages, including, for each linkage that was sought, whether the linking process was described and the success rate was reported, and, if applicable, whether information had been obtained from maternal or infant files; and (4) analytical aspects, including the explicit mention of the unit of analysis for pregnancy (e.g., pregnancy, unique woman) and fetal or infant outcomes (e.g., fetus, child, singleton pregnancy), the gestational age at start of follow-up (e.g., median gestational age at first contact with the health care system, mean gestational age at enrollment), and whether intrafamily correlation had been considered in the study design or analysis (e.g., restricting the study population to one pregnancy per woman, implementing robust variance analyses).

### 2.2 Literature search and article selection

This literature search was designed to identify observational studies on the utilization or safety of medical products during pregnancy using any type of data source. Safety studies could have any type of comparator population, including historical comparison groups, and could have pregnancy, maternal, neonatal, or pediatric outcomes. No limits were established based on geographical region; publications had to be written in English and have a PubMed publication date in 2015-2018. Animal studies were excluded. The search was structured using the PICOTS (population, intervention, comparison, outcomes, time, setting) framework^5^ (Appendix A, Table A-1) and was applied in PubMed (search terms are presented in Appendix A, Table A-2).

Eligibility criteria were applied during screening of titles or titles and abstracts (level 1 screening, conducted by one researcher) and screening of full-text articles (level 2 screening, conducted by one researcher). Briefly, publications had to have an abstract and present original research; exposure groups had to be determined based on use of medical products (with or without the diagnosis of a condition). Detailed eligibility criteria are provided in Appendix A, Table A-3. Publications could have more than one reason for exclusion, but only one was selected hierarchically (i.e., publication type, exposure, outcomes, language, and population) to facilitate assessment of each publication’s disposition. See Appendix A, Table A-4, for exclusion criteria. Doubts on inclusion were solved by discussion among the authors.

For practical reasons, this literature review had a target size of 100 publications. To restrict the number of publications, the following process was implemented: (1) all retrieved records underwent level 1 screening; (2) each publication that passed level 1 screening and was available for level 2 screening was assigned a random number with a uniform distribution between 0 and 1, and publications were sorted from smallest to largest on this number; and (3) publications underwent level 2 screening in this order until reaching the target size of 100 included publications.

### 2.3 Data extraction and reporting of key methodological elements

Data were extracted by one researcher and quality checked against the original source by a second researcher. Data extracted included study characteristics (e.g., study size, study design, whether the study had been requested by a regulatory agency, and whether it was published in an epidemiology or pharmacoepidemiology journal or in a journal with a different focus) and whether the key methodological elements were explicitly reported in the publication. For example, for the element “Has the intrafamily correlation been considered?” the data extractor recorded “yes” if the publication mentioned that only one sibling per family was included, that robust variance was used to account for sibling correlation, or if the study was a sibling design, among other possible strategies. If the publication did not include information on the topic, the data extractor recorded “no.” Some methodological elements were not applicable to a given study (e.g., in studies of stillbirth, linkage to infant files is not possible); additional criteria are listed in Appendix B, Notes tab. When a publication reported on more than one population or analysis, the data extractor recorded “yes” if the information on a given element was reported for at least one of the populations or analyses. Other specifications for data extraction can be found in Appendix B, Look-ups and Notes tabs. We calculated the prevalence of reporting of each key element as a percentage in which the denominator was the number of publications for which the element was applicable, and the numerator was the number of publications that reported on that element.

## 3 RESULTS

### 3.1 Literature search and selected articles

Our search identified 1,981 unique articles for level 1 screening, of which 406 progressed to level 2 screening (Figure 1). The target size of 100 publications was achieved after screening the full text of 139 publications.

**Figure 1.**
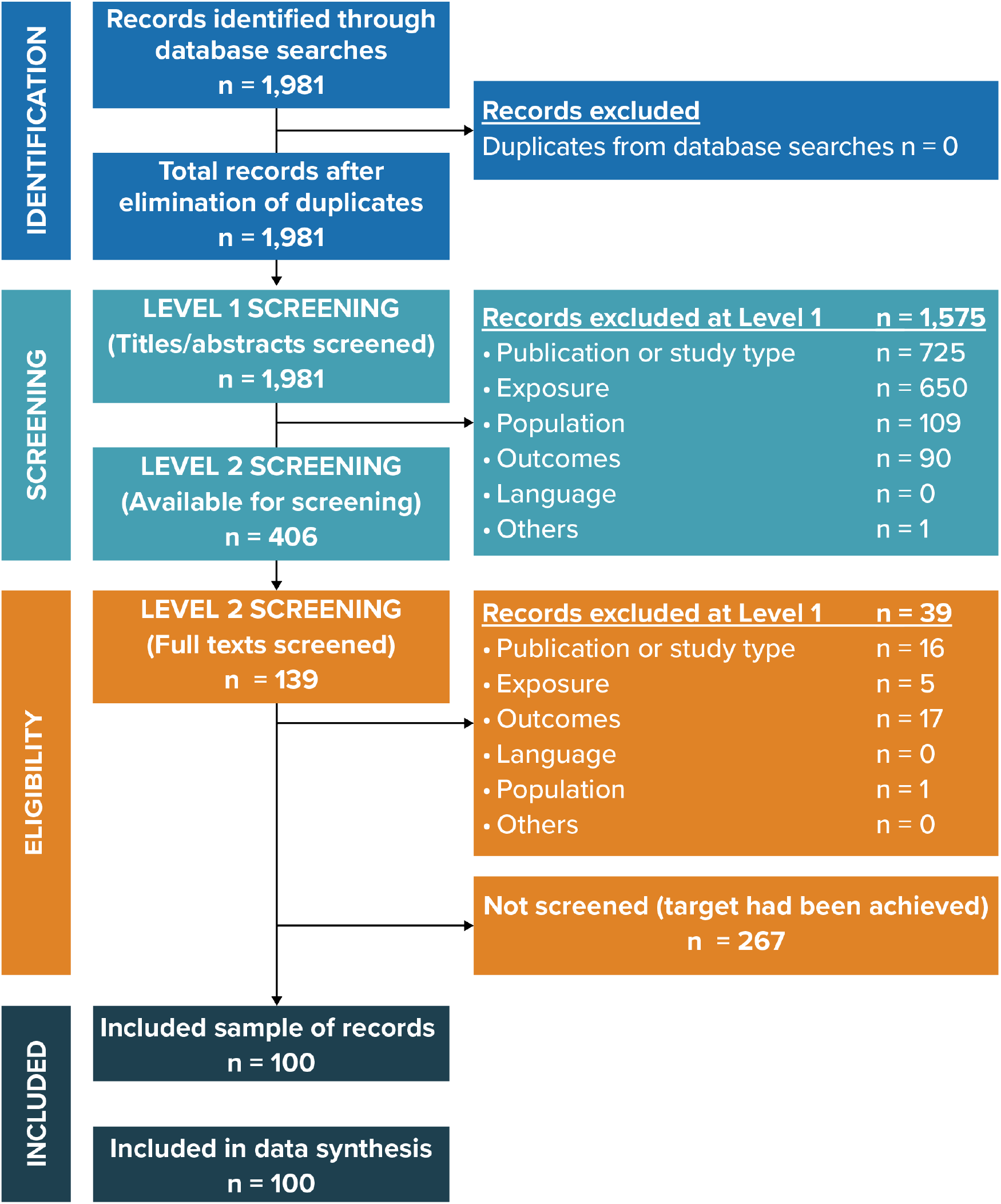
Screening and selection of articles into the literature review. Note: This is a PRISMA chart.^33^ Abbreviation: PRISMA, Preferred Reporting Items for Systematic Reviews and Meta-Analyses.

The number of pregnancy pharmacoepidemiology publications increased over time: of the 100 publications in this review, 17% were published in 2015, 22% in 2016, 22% in 2017, 35% in 2018, and 4% in 2019 (Appendix B, Data extraction tab). Our search had a publication date limit of December 31, 2018; however, the publication year of the actual publication was not always consistent with the publication date in PubMed records (possibly reflecting early online vs. final publication date in some instances). Of publications in this review, 8% were published in journals with a focus on epidemiology or pharmacoepidemiology and 92% were reported in general medical journals, journals focusing on a medical specialty, or journals with another focus. No publication reported that the study had been conducted to fulfill a request from a regulatory agency, but one was conducted as part of an evaluation of a state vaccination program.^6^ Of the 100 publications, 14% were for medical product–utilization studies; 86% had a medical product–safety component. Data sources used were health care claims data (14% of publications), electronic medical records (14%), paper medical records (9%), health care registries from the Nordic countries (8%), and pregnancy exposure registries (3%); the remainder used other data sources. The median study size was 2025 women, pregnancies, or offspring (mean, 126,843).

### 3.2 Reporting of key methodological elements

Denominators for percentages reported in this section vary, as some elements were applicable to only a subset of the 100 included studies (Tables 1-4).

**Table 1.**
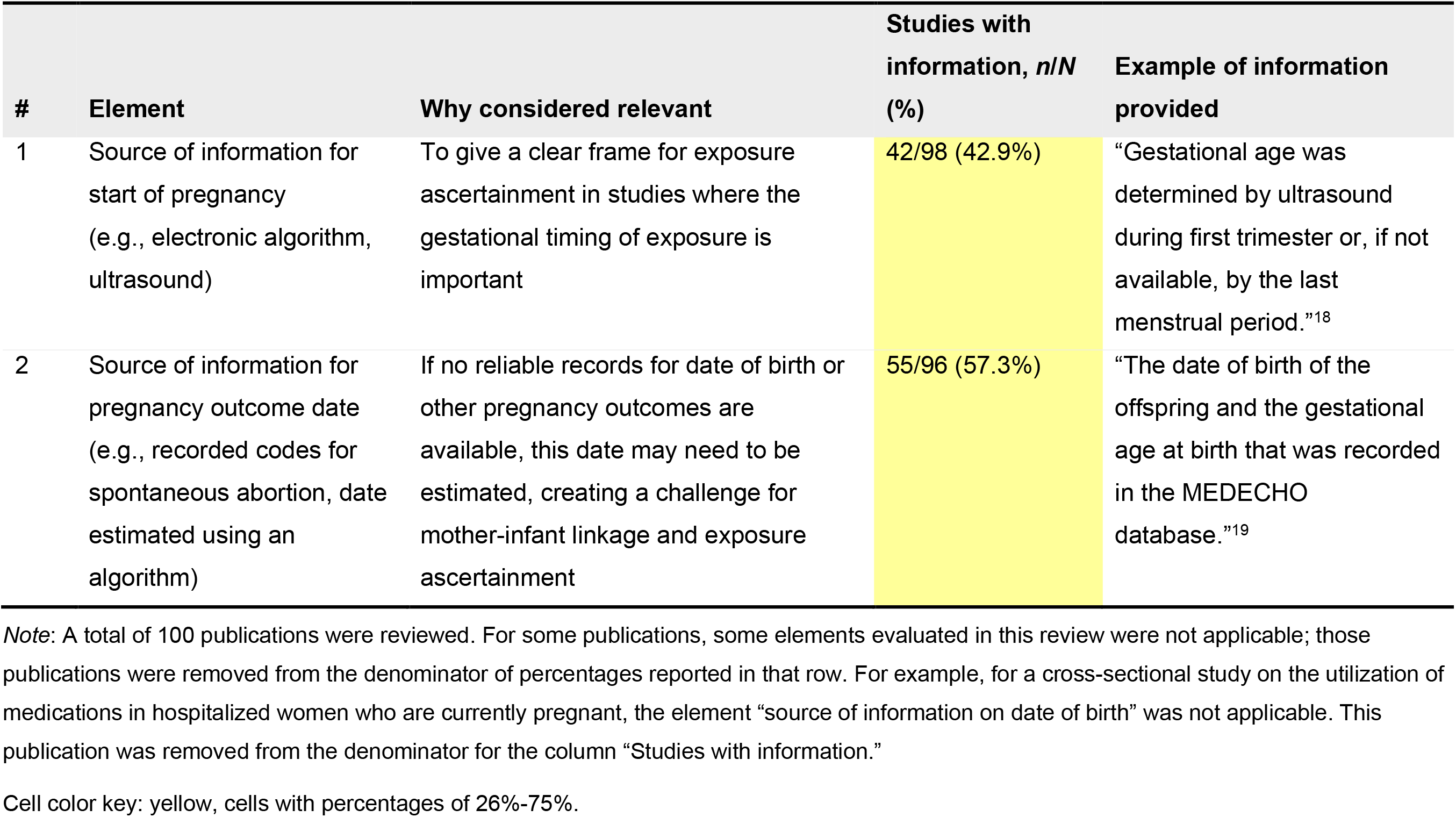
Source of information on beginning and end of pregnancy.

| # | Element | Why considered relevant | Studies with information, *n/N* (%) | Example of information provided |
| --- | --- | --- | --- | --- |
| 1 | Source of information for start of pregnancy (e.g., electronic algorithm, ultrasound) | To give a clear frame for exposure ascertainment in studies where the gestational timing of exposure is important | 42/98 (42.9%) | "Gestational age was determined by ultrasound during first trimester or, if not available, by the last menstrual period."<sup>18</sup> |
| 2 | Source of information for pregnancy outcome date (e.g., recorded codes for spontaneous abortion, date estimated using an algorithm) | If no reliable records for date of birth or other pregnancy outcomes are available, this date may need to be estimated, creating a challenge for mother-infant linkage and exposure ascertainment | 55/96 (57.3%) | "The date of birth of the offspring and the gestational age at birth that was recorded in the MEDECHO database."<sup>19</sup> |

*Note:* A total of 100 publications were reviewed. For some publications, some elements evaluated in this review were not applicable; those publications were removed from the denominator of percentages reported in that row. For example, for a cross-sectional study on the utilization of medications in hospitalized women who are currently pregnant, the element "source of information on date of birth" was not applicable. This publication was removed from the denominator for the column "Studies with information."
Cell color key: yellow, cells with percentages of 26%-75%.

#### Source of information on start and end of pregnancy

Of the publications for which these elements were applicable, 43% reported on the source for determining the start of pregnancy, and 57% reported on the source for determining the end of pregnancy (Table 1).

#### Composition of the study population

Reporting in this domain ranged from 27% of publications reporting on whether fetuses with chromosomal abnormalities were included in the study population to 93% reporting on whether non-live births were included (Table 2). Of publications included, 57% reported whether multifetal pregnancies were included in the study population, 39% reported whether more than one pregnancy was included in the study population, 60% reported whether fetuses with major malformations were included, and 36% reported whether fetuses with minor malformations were included.

**Table 2.** Composition of the study population.

| # | Element | Why considered relevant | Studies with information, n/N (%) | Example of information provided |
| --- | --- | --- | --- | --- |
| 3 | Multifetal pregnancies included in study population? | To assess the potential for intrafamily correlation<br>To provide context on whether the study population is at greater risk for outcomes that are known to be associated with multifetal pregnancies | 55/97 (56.7%) | "All pregnancies of single and twin births were considered."<sup>20</sup> |
| 4 | More than one pregnancy per woman included in study population? | To understand the composition of the study population<br>To identify the potential for intrafamily correlation | 36/93 (38.7%) | "...131 women (144 pregnancies) were exposed..."<sup>15</sup> |
| 5 | Fetuses with chromosomal abnormalities included in study population? | To assess the potential for recall bias in self-reported exposure<br>To understand the outcome definition in research on congenital malformations (most infants with chromosomal abnormalities have congenital malformations, some of which might be related to the chromosomal abnormality) | 24/88 (27.3%) | "Birth defects include genetic syndromes and chromosomal abnormalities, but both these types of abnormalities were excluded from the calculation of the birth defect rates."<sup>21</sup> |
| 6 | Fetuses with major malformations included in study population? | To assess the potential for recall bias in self-reported exposure<br>To put in context results from analyses of outcomes that might be affected in the presence of major malformations, such as infant's size at birth | 53/88 (60.2%) | "We analyzed...among non-malformed singleton controls in the National Birth Defects Prevention Study."<sup>22</sup> |
| 7 | Fetuses with minor malformations included in study population? | To understand whether fetuses with minor malformations were considered noncases in research on major congenital malformations | 32/88 (36.4%) | "All observed major and minor birth defects..."<sup>23</sup> |
| 8 | Non-live births included in denominator? | To assess any potential for bias due to not including in the population all fetuses at risk | 76/82 (92.7%) | "In this population based study we included women...who gave birth to a live singleton infant..."<sup>24</sup> |

*Note:* A total of 100 publications were reviewed. For some publications, some elements evaluated in this review were not applicable; those publications were removed from the denominator of percentages reported in that row. For example, for a cross-sectional study on the utilization of medications in hospitalized women who are currently pregnant, the element "source of information on date of birth" was not applicable. This publication was removed from the denominator for the column "Studies with information."
Cell color key: yellow, cells with percentages of 26%-75%; green, 76%-100%.

#### Mother-infant and father-infant linkages

Mother-infant linkage was sought in 20 publications; 35% of the publications described the process and 21% reported the success rate (Table 3). Of the 20 publications, 65% specified which information had been obtained from maternal records and which information had been obtained from infant records. Father-infant linkage was sought by two studies in this sample; one described the process and the other reported the linkage success rate.

**Table 3.**
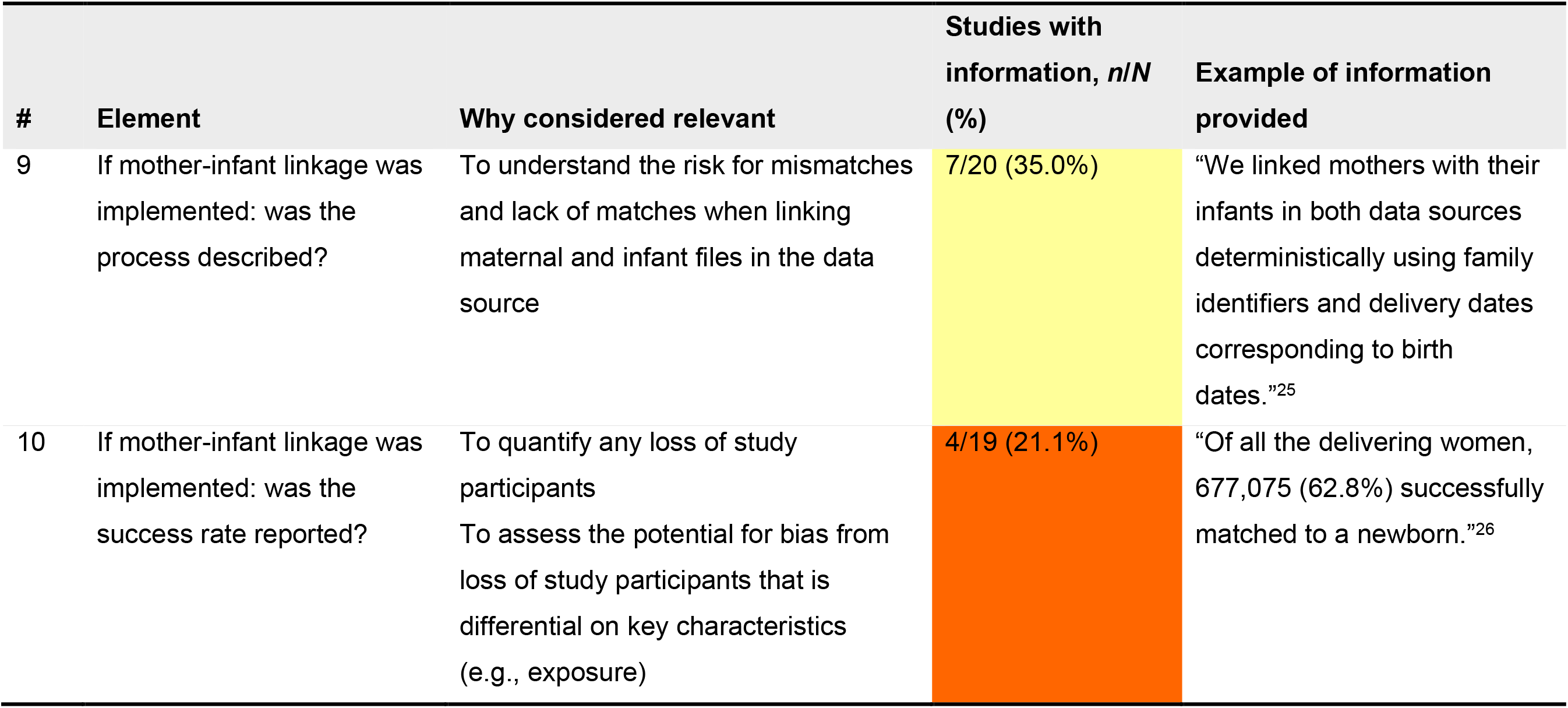
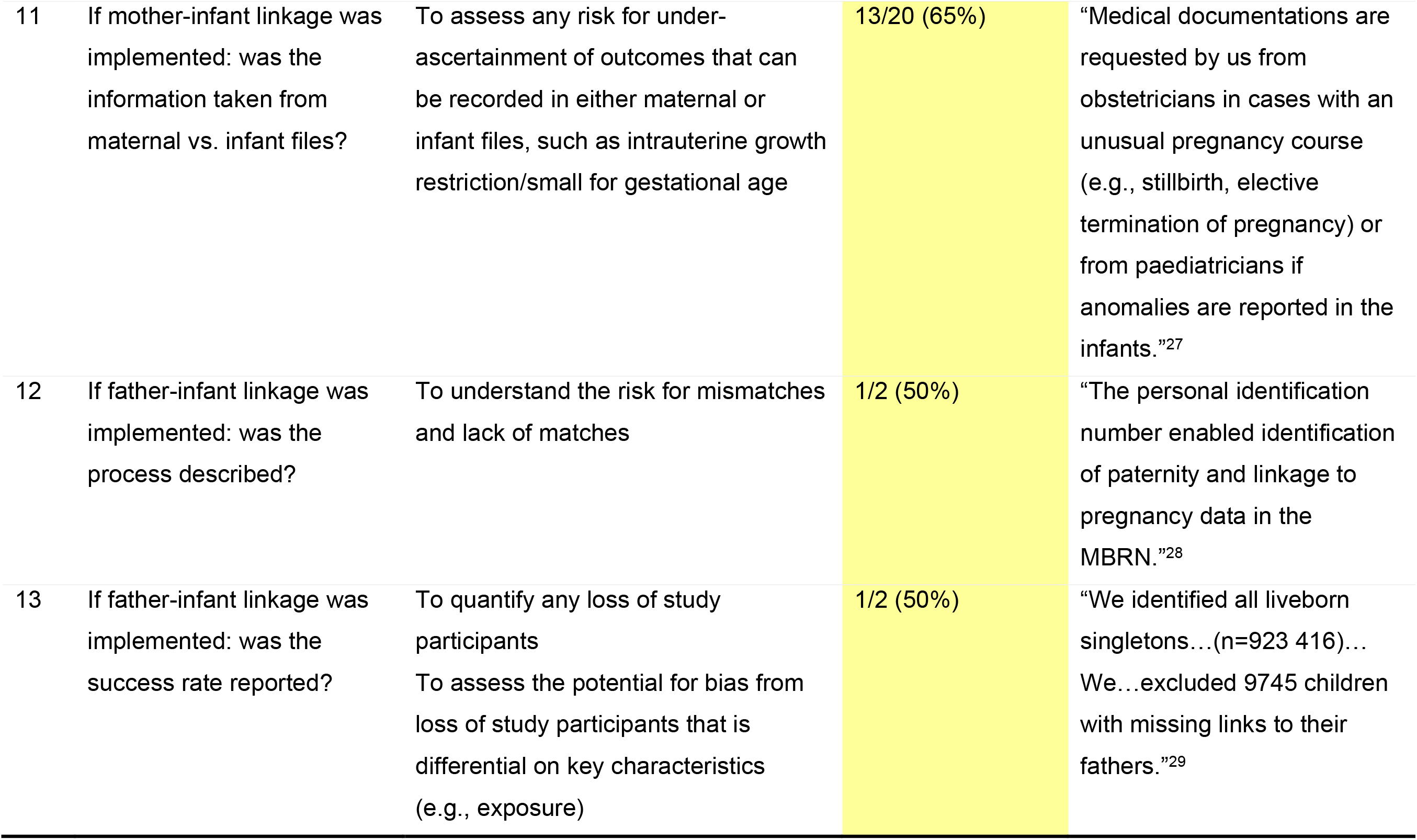
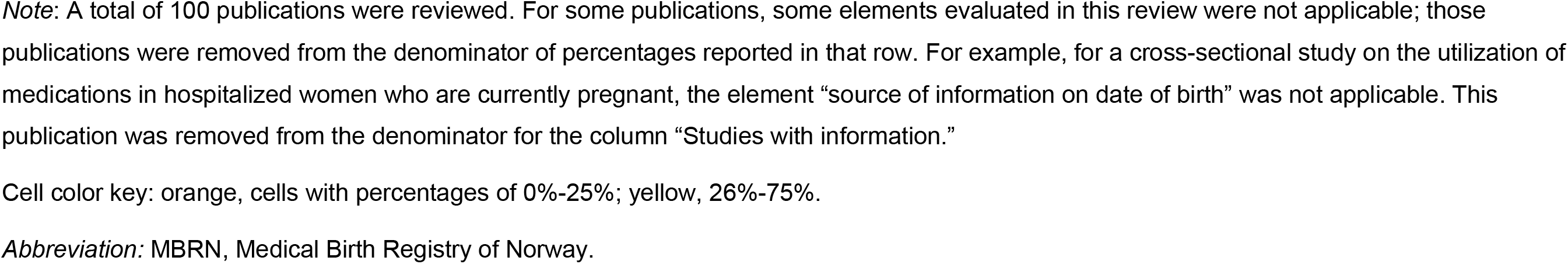
Mother-infant and father-infant linkages.

#### Analytical aspects

The unit of analysis was reported in 98% of the publications with pregnancy outcomes and in 94% of the publications with fetal or infant outcomes (Table 4). Gestational age at start of follow-up was reported by 43% of publications, and whether the intrafamily correlation was considered in analysis or by design was reported by 22% of publications.

**Table 4.**
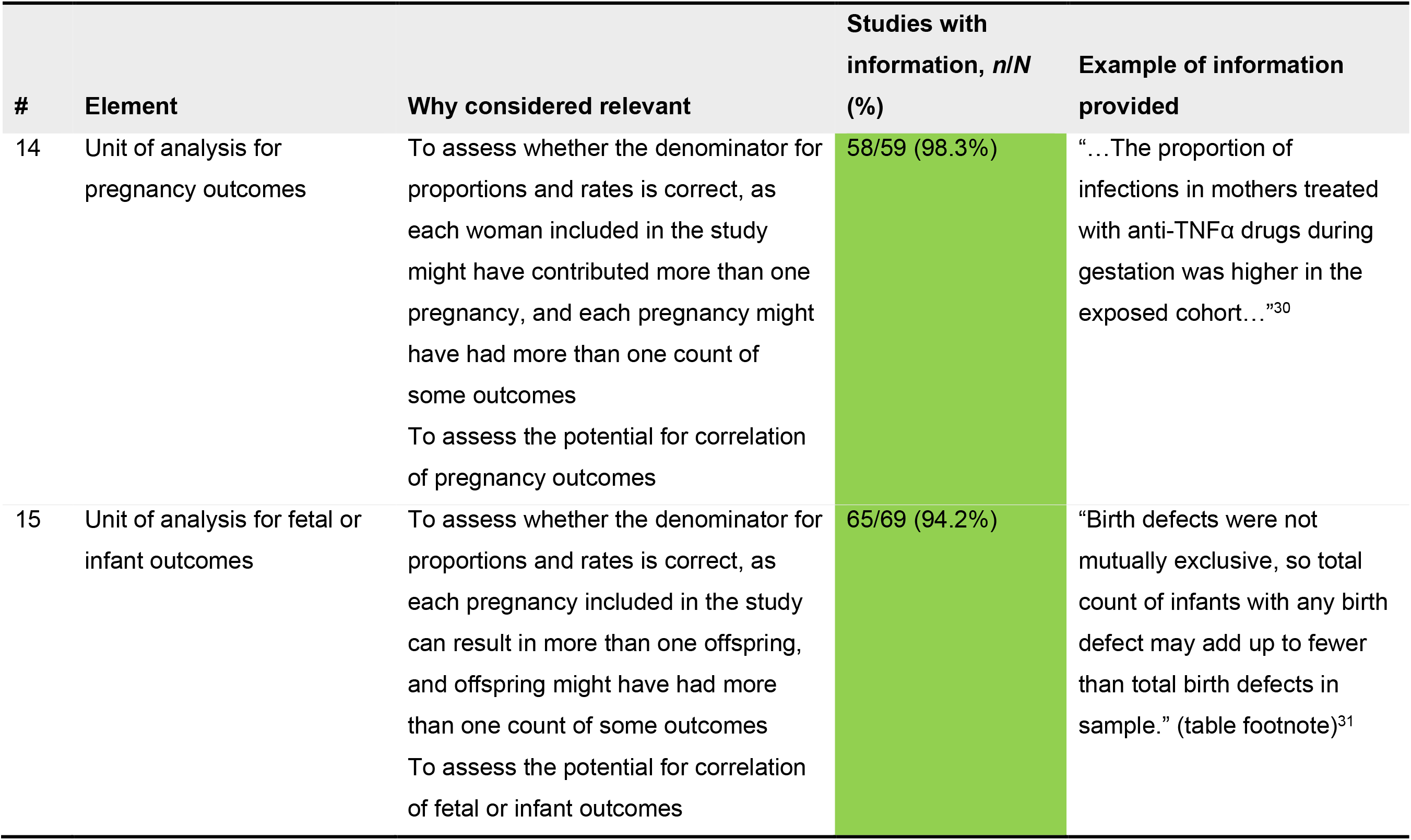
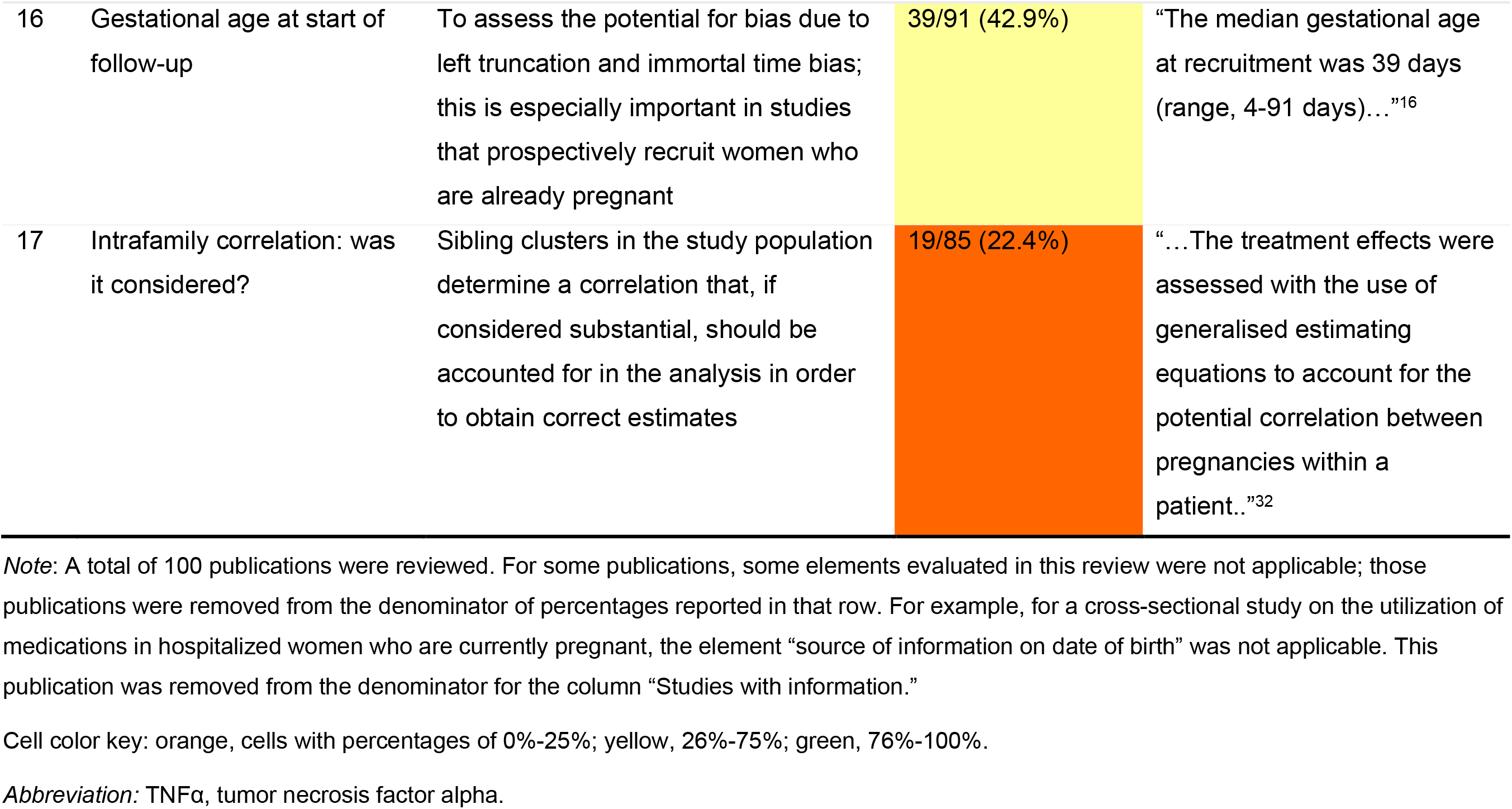
Analytical aspects.

## 4 DISCUSSION

To our knowledge, no published consensus exists on which key methodological elements should be reported in studies on the utilization or safety of medical products in pregnancy. Based on regulatory guidelines, the literature, and subject-matter knowledge, we developed a list of key methodological elements for pregnancy pharmacoepidemiology that includes elements related to the source of information on start and end of pregnancy, composition of the study population, mother-infant and father-infant linkages, and analytical aspects. In a sample of 100 publications on pregnancy pharmacoepidemiology published in 2015-2019, we observed that completeness of reporting was heterogeneous across these key elements. At one end of the spectrum, nearly all publications reported the unit of analysis for pregnancy outcomes or whether non-live births were included in the denominator of calculations. At the other end, only about one fifth of publications reported on whether intrafamily correlation had been considered or the success of mother-infant linkage. Father-infant linkage was sought in only two publications.

We propose that a concise checklist for pregnancy pharmacoepidemiology studies might help improve the reporting of these key elements. A checklist could be structured like a section of the ENCePP Checklist for Study Protocols,^7^ with tick boxes for “yes,” “no,” and “not applicable” and a cell to specify the document section in which the information is provided. Such a checklist could be applied to study protocols and publications.

Some of the elements that we identified are not applicable to all studies. For example, the source of information for pregnancy outcomes is irrelevant in a cross-sectional study that describes current use of medications in hospitalized women who are currently pregnant.^8^ We did not consider that reporting on mother-infant linkage was applicable to hospital-based studies or to studies that derived their data from questionnaires. In general, though, we believe that for studies that link maternal and infant records (or records from other family members), the process and success rate should be reported, even if the process is simple and the success rate is expected to be close to 100%, as may be the case with countrywide Nordic health care registries. This is because some readers may be unfamiliar with the data sources used in a study, and processes may change over time. Also, reference to a previously published paper does not help a reader who may not have immediate access to that publication. Although we did not assess completeness of reporting in relation to linkage to registries of major congenital malformations,^9^ with vital statistics records,^10^ or other data sources, ideally, the linkage methods and success rates should be reported.

None of the sampled publications reported that the underlying study had been conducted to meet regulatory requirements, although our sample included pregnancy exposure registries that are generally established to meet such requirements. One study was conducted as part of an evaluation of a state vaccination program.^6^ For transparency and to provide context for the study design, we recommend that information on whether studies have been conducted to meet regulatory requirements be included in scientific publications.

We noticed ambiguity in the use of “multiple pregnancies,” which was used to refer to multifetal pregnancies^11,12^but also to more than one (singleton or multifetal) pregnancy in the same woman.^13-15^ There is no ambiguity in obstetrical practice, where one pregnancy per woman is evaluated at a given time, but ambiguity can arise in studies where each woman can contribute with more than one pregnancy during her longitudinal follow-up. Although the context generally provides clarity on the intended meaning, we recommend avoiding ambiguous language.

Limitations of this review include the relatively small sample of publications included in the literature review. Although the results presented here may not be generalizable to the entire body of publications on pregnancy pharmacoepidemiology research, they can support recommendations regarding completeness of reporting.

A degree of subjectivity was involved in deciding whether some elements were applicable to a given study. To address this, the authors developed internal guidance in relation to specific types of data sources (presented in Appendix B, Notes tab) and decided to assume that, for consistency, when in doubt, they would consider that the element was applicable. For this reason, this review may have overestimated the extent to which the elements were not reported.

Furthermore, authors of included studies may not have reported some elements in our list because they may not have considered these elements to be relevant for their study. This might have been true for the element “whether pregnancies carrying fetuses with chromosomal abnormalities or malformations were included in the study population.” For example, for studies in which the exposure or the outcomes were not obviously related to chromosomal abnormalities or malformations, or in drug utilization studies, one might assume that such pregnancies were included in the study without an explicit mention. In other publications, authors may have reported that infants with major malformations were excluded, implying that infants with minor malformations were included. We believe that more thorough reporting would bring more clarity than harm.

In addition, we observed variation in the amount of information provided on gestational age at the start of follow-up. For example, “The median gestational age at recruitment was 39 days (range, 4-91 days)…”^16^ is more specific than “Pregnant women who were diagnosed…at 16-20 weeks’ gestation.”^17^ However, both contain information on gestational age at enrollment.

This review ascertained key methodological elements that are specific to pregnancy pharmacoepidemiology. Other elements, common with observational studies in other areas, were not assessed; this was done to avoid overlap with existing checklists and to keep our list of elements as focused as possible.

In conclusion, completeness of reporting the methods used in pregnancy pharmacoepidemiology studies can be improved. This would facilitate the interpretation of study results and the comparison of results across studies. A purpose-made checklist would help to increase transparency in the dissemination of results of studies on utilization or safety of medical products in pregnancy.

## Data Availability

The data are contained within the manuscript and its appendices.

## ETHICS STATEMENT

All authors are employees of RTI Health Solutions, a unit of RTI International, which is an independent, not-for-profit organization that conducts work for government, public, and private organizations including pharmaceutical companies. The authors do not have any conflicts of interest for this work.

## ACKNOWLEDGEMENTS

Editorial services were provided by Adele Monroe, and graphic art services were provided by Bethan Pickering, both employees of RTI Health Solutions. We would like to thank Abenah Harding, also an employee of RTI Health Solutions, for her help preparing this manuscript. Development of this manuscript was supported financially by RTI Health Solutions.

## Appendix A. Study eligibility criteria and literature search terms

This appendix contains the study eligibility criteria and search terms for the literature search and review.

**Table A-1.** Study eligibility defined per PICOTS and additional criteria.

| Aspect | Description |
| --- | --- |
| Study type | Observational study on safety or utilization of medical products in pregnancy<br>Original research |
| Language | English |
| Patient population | Pregnant women |
| Intervention (exposure) | Any drug/vaccine/biologic/device (i.e., medical product) used in pregnancy |
| Comparator | Any comparator, including historical comparators, or no comparators in utilization studies |
| Outcomes | Pregnancy, maternal, neonatal, or pediatric outcomes, or no outcome in utilization studies |
| Timing | Duration of treatment: any<br>Duration of follow-up: any<br>Publication date: 2015 to 2018 |
| Setting | Any (e.g., ambulatory, in-hospital) |

Abbreviation: PICOTS, population, intervention, comparison, outcomes, time, setting.<sup>1</sup>

**Table A-2.**
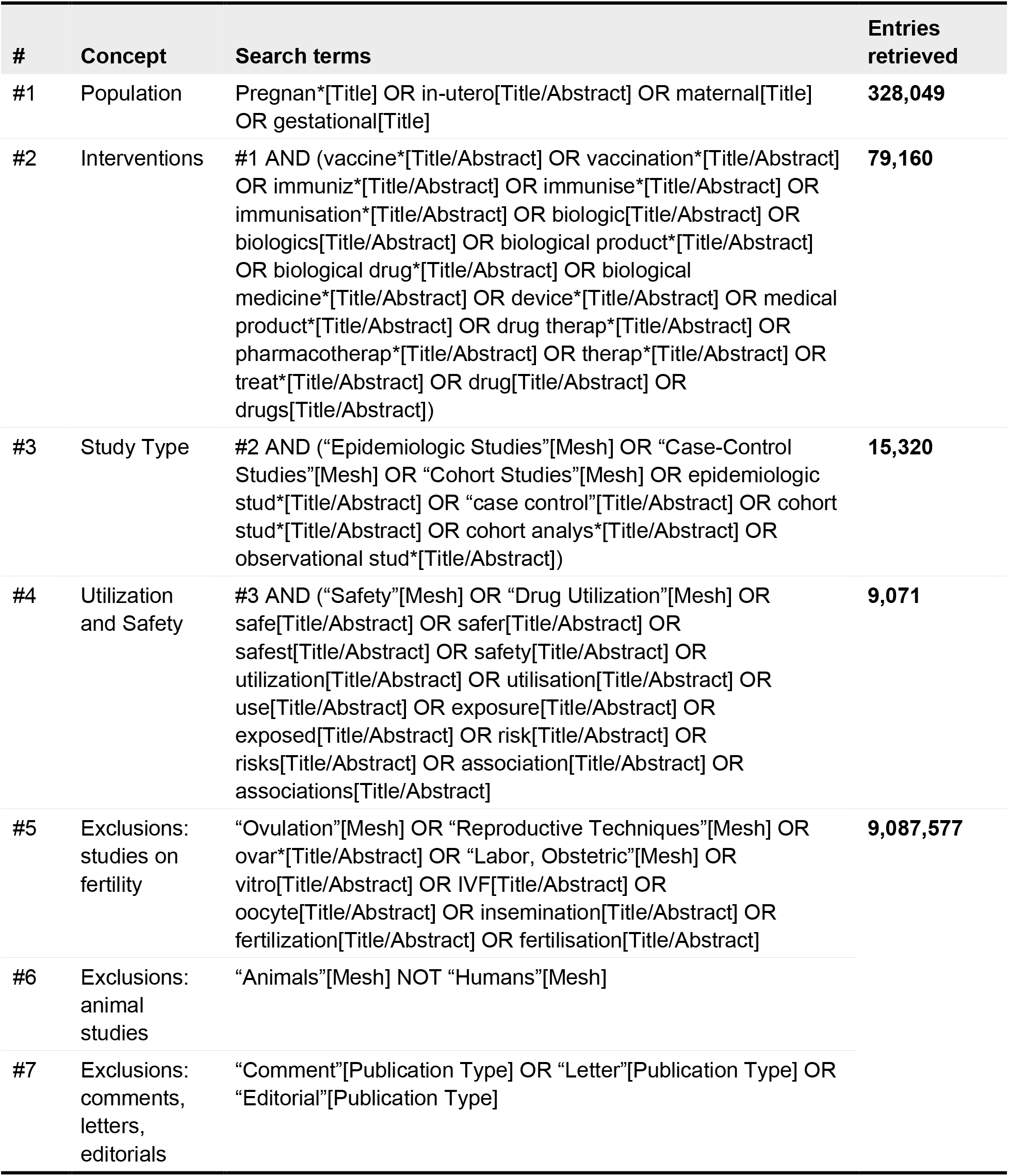
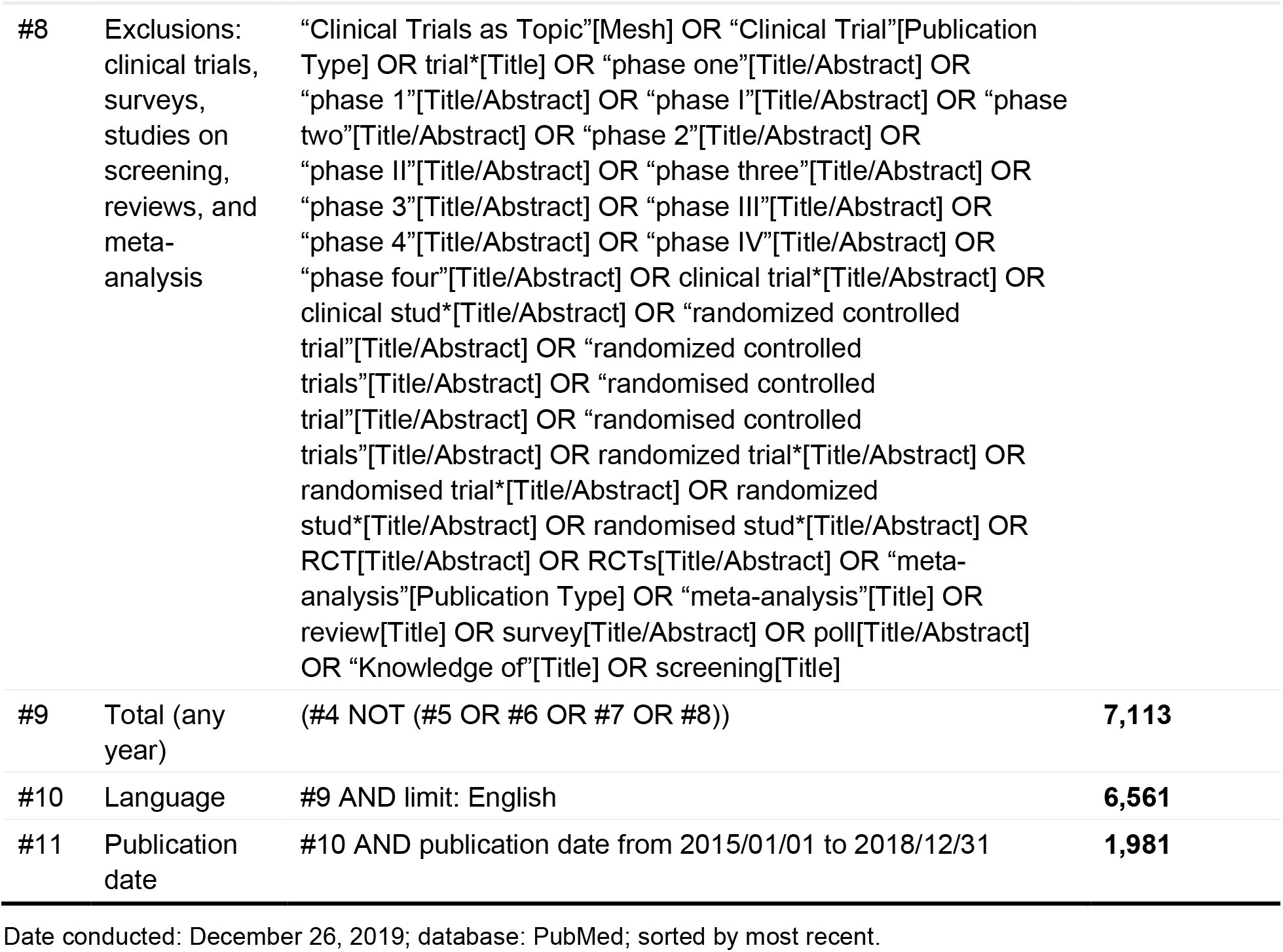
Search terms and results.

| # | Concept | Search terms | Entries retrieved |
| --- | --- | --- | --- |
| #1 | Population | Pregnan*[Title] OR in-utero[Title/Abstract] OR maternal[Title] OR gestational[Title] | **328,049** |
| #2 | Interventions | #1 AND (vaccine*[Title/Abstract] OR vaccination*[Title/Abstract] OR immuniz*[Title/Abstract] OR immunise*[Title/Abstract] OR immunisation*[Title/Abstract] OR biologic[Title/Abstract] OR biologics[Title/Abstract] OR biological product*[Title/Abstract] OR biological drug*[Title/Abstract] OR biological medicine*[Title/Abstract] OR device*[Title/Abstract] OR medical product*[Title/Abstract] OR drug therap*[Title/Abstract] OR pharmacotherap*[Title/Abstract] OR therap*[Title/Abstract] OR treat*[Title/Abstract] OR drug[Title/Abstract] OR drugs[Title/Abstract]) | **79,160** |
| #3 | Study Type | #2 AND ("Epidemiologic Studies"[Mesh] OR "Case-Control Studies"[Mesh] OR "Cohort Studies"[Mesh] OR epidemiologic stud*[Title/Abstract] OR "case control"[Title/Abstract] OR cohort stud*[Title/Abstract] OR cohort analys*[Title/Abstract] OR observational stud*[Title/Abstract]) | **15,320** |
| #4 | Utilization and Safety | #3 AND ("Safety"[Mesh] OR "Drug Utilization"[Mesh] OR safe[Title/Abstract] OR safer[Title/Abstract] OR safest[Title/Abstract] OR safety[Title/Abstract] OR utilization[Title/Abstract] OR utilisation[Title/Abstract] OR use[Title/Abstract] OR exposure[Title/Abstract] OR exposed[Title/Abstract] OR risk[Title/Abstract] OR risks[Title/Abstract] OR association[Title/Abstract] OR associations[Title/Abstract]) | **9,071** |
| #5 | Exclusions: studies on fertility | "Ovulation"[Mesh] OR "Reproductive Techniques"[Mesh] OR ovar*[Title/Abstract] OR "Labor, Obstetric"[Mesh] OR vitro[Title/Abstract] OR IVF[Title/Abstract] OR oocyte[Title/Abstract] OR insemination[Title/Abstract] OR fertilization[Title/Abstract] OR fertilisation[Title/Abstract] | **9,087,577** |
| #6 | Exclusions: animal studies | "Animals"[Mesh] NOT "Humans"[Mesh] |  |
| #7 | Exclusions: comments, letters, editorials | "Comment"[Publication Type] OR "Letter"[Publication Type] OR "Editorial"[Publication Type] |  |
| #8 | Exclusions: clinical trials, surveys, studies on screening, reviews, and meta-analysis | “Clinical Trials as Topic”[Mesh] OR “Clinical Trial”[Publication Type] OR trial*[Title] OR “phase one”[Title/Abstract] OR “phase 1”[Title/Abstract] OR “phase I”[Title/Abstract] OR “phase two”[Title/Abstract] OR “phase 2”[Title/Abstract] OR “phase II”[Title/Abstract] OR “phase three”[Title/Abstract] OR “phase 3”[Title/Abstract] OR “phase III”[Title/Abstract] OR “phase 4”[Title/Abstract] OR “phase IV”[Title/Abstract] OR “phase four”[Title/Abstract] OR clinical trial*[Title/Abstract] OR clinical stud*[Title/Abstract] OR “randomized controlled trial”[Title/Abstract] OR “randomized controlled trials”[Title/Abstract] OR “randomised controlled trial”[Title/Abstract] OR “randomised controlled trials”[Title/Abstract] OR randomized trial*[Title/Abstract] OR randomised trial*[Title/Abstract] OR randomized stud*[Title/Abstract] OR randomised stud*[Title/Abstract] OR RCT[Title/Abstract] OR RCTs[Title/Abstract] OR “meta-analysis”[Publication Type] OR “meta-analysis”[Title] OR review[Title] OR survey[Title/Abstract] OR poll[Title/Abstract] OR “Knowledge of”[Title] OR screening[Title] |  |
| #9 | Total (any year) | (#4 NOT (#5 OR #6 OR #7 OR #8)) | 7,113 |
| #10 | Language | #9 AND limit: English | 6,561 |
| #11 | Publication date | #10 AND publication date from 2015/01/01 to 2018/12/31 | 1,981 |

Date conducted: December 26, 2019; database: PubMed; sorted by most recent.

**Table A-3.** Study eligibility criteria.

| Study eligibility criteria |
| --- |
| ▪ The publication has an abstract<br>▪ The abstract and the full-text publication are written in English<br>▪ The publication presents original observational research that focuses on the utilization or safety of a drug/vaccine/biologic/device in pregnancy and presents numerical results (i.e., study protocols are not eligible; studies on surgery or procedures are not eligible)<br>▪ Exposure groups (i.e., the exposed and the unexposed) are based on the use of medical products in pregnancy (with or without a diagnosis of a condition); studies that determine exposure groups based on diagnoses only are not eligible<br>▪ Studies using any type of data source are eligible<br>▪ Additional criteria for level 2 screening: none |

**Table A-4.** Reasons for study exclusion during screening.

| Hierarchy for exclusion of studies |
| --- |
| ▪ Publication type: studies that are not original research, are not observational, or do not focus on drug/vaccine/biologic/device utilization or safety in pregnancy<br>▪ Exposure: studies that do not report on a drug/vaccine/biologic/device (e.g., studies that report on epilepsy rather than on antiepileptics)<br>▪ Outcomes: studies that do not focus on the utilization or safety of these products (e.g., studies on efficacy of drugs)<br>▪ Language: studies not written in English<br>▪ Population: studies that are not on pregnant women (e.g., studies on animals or studies on women who are not pregnant) |

